# Choline and betaine concentrations in plasma predict dietary choline intake in healthy humans: a double-blind randomized control feeding study

**DOI:** 10.1101/2025.07.15.25331567

**Authors:** Isis Trujillo-Gonzalez, David A. Horita, Julie Stegall, Rachel Coble, Evan M. Paules, Anju A. Lulla, Emmanuel Baah, Teodoro Bottiglieri, Wei Sha, Martin Kohlmeier, Walter B. Friday, Steven H. Zeisel

## Abstract

**Background:** Choline is an essential nutrient, and insufficient intake negatively affects organs such as the liver, brain, and muscles. In the United States, average choline intake remains below the Adequate Intake (AI) (550 mg/day men, 425 mg/day women). Although conventional dietary assessment tools can identify people who are eating diets low in choline, no metabolite biomarkers have been proven to reliably assess choline intake.

**Objective:** We tested whether plasma concentrations of choline and its metabolites could determine dietary choline intake. We also assessed whether liver elastography (Fibroscan) could detect diet-induced changes in liver fat.

**Methods:** In a double-blind, randomized, crossover feeding study, participants adhered to 3 distinct dietary patterns for 2-wk intervals, delivering approximately 100%, 50%, and 25% of the choline AI. On Day 12 of each dietary arm, in addition to the food supplied, subjects consumed a single bolus of 2.2 mmol trimethyl-d_9_-choline. Plasma concentrations of choline, betaine and phosphatidylcholine (PtdCho) were measured using mass spectrometry. Targeted assays quantified choline, betaine, phosphatidylcholine and total homocysteine concentrations. Liver fat content was evaluated non-invasively using Fibroscan.

**Results:** Plasma concentrations of d_9_-choline, betaine, and their isotopic enrichment ratio (IER) varied with dietary intake (q<0.0001), and PtdCho IER also differed significantly (q=0.001). In targeted analysis, choline and betaine concentrations were highly responsive to dietary choline intake, while PtdCho and tHcy were not. Receiver Operator Characteristic (ROC) analysis showed strong accuracy using plasma choline (AUC=0.81) and betaine (AUC=0.80), with improved accuracy when combined (AUC= 0.83). Fibroscan identified a subset of participants with increased liver fat in response to the 25% AI vs. 100% AI choline diet, though patterns varied among individuals.

**Conclusion:** Plasma choline and betaine concentrations are robust biomarkers of dietary choline intake under controlled feeding. These findings support targeted metabolite profiling to improve choline intake assessment and reveal induvial variability in liver response to low choline intake.

## INTRODUCTION

Choline is an essential nutrient that serves as a precursor for several metabolites critical for human health. These metabolites include phosphatidylcholine (PtdCho), a major structural component of cell membranes; acetylcholine (ACho), a neurotransmitter; and betaine, a methyl donor involved in the regulation of gene expression (1, 2). While choline can be synthesized *de novo* from phosphatidylethanolamine via the enzyme phosphatidylethanolamine-*N*-methyltransferase (PEMT), endogenous synthesis alone is insufficient to meet daily choline requirements for most people. Therefore, dietary intake of choline is necessary to maintain optimal health. Low dietary choline intake in adults is associated with adverse effects such as hepatic fat accumulation and muscle dysfunction (3, 4); and underconsumption of choline during pregnancy increases the risk of having a baby with neural tube defects (5).

The adequate intake (AI) levels for choline were established by the Institute of Medicine (now the National Academy of Medicine) as part of the Dietary Reference Intakes in 1998 (6). The recommended intake is 550 mg/day for men and 425 mg/day for women, increasing to 450 mg/ day during pregnancy and lactation (2). However, data from the 2013-2016 NHANES survey in the United States indicate that most individuals do not meet these recommendations (7, 8). In addition, studies using food frequency questionnaires have highlighted a widespread trend of low choline dietary intake among women of reproductive age. For example, in The Gambia intake in premenopausal women was estimated at just 155/mg per day (9). In Mexico and Taiwan, reported intakes are approximately 262 mg/day and 284 mg/ day, respectively (10, 11). However, the accuracy of these dietary assessments is limited by the inherent weaknesses of food frequency questionnaires, including imprecise questions, lack of detail on food items, and underreporting. These limitations raise concerns about the reliability of using such data to assess choline intake (12).

Genetic variation adds a layer of complexity to choline metabolism. Single nucleotide polymorphisms (SNPs), such as rs12325817, located in the promoter region of the *PEMT* gene, can reduce gene expression and thereby increase reliance on dietary choline (13). Moreover, the promoter region of the *PEMT* gene is regulated by estrogen-responsive elements (EREs) (14, 15), which partly explains why premenopausal women consuming a low-choline diet are less likely to experience organ dysfunctions associated with low choline intake compared to menopausal women and men (16). In controlled feeding studies, choline deprivation in men led to fatty liver, elevated plasma levels of hepatic enzymes such as aspartate aminotransferase (AST) and alanine aminotransferase (ALT), and increased muscle breakdown indicated by elevated plasma creatine kinase (CK) (4, 17). Although choline is recognized as an essential nutrient and recent advances have shed light on choline transport (18, 19), our ability to accurately measure dietary choline intake in human populations remains limited. Given the central role of choline and its metabolites in maintaining metabolic and organ health, this study evaluated multiple approaches to estimate dietary choline intake before clinical manifestations occur, identifying plasma metabolites as the most informative markers.

## METHODS

### Study design

The study design was a longitudinal, double-blind, randomized crossover dietary intervention (ClinicalTrials.gov identifier: NCT03726671). Choline content of the dietary intervention was based on the National Institute of Medicine-defined Adequate Intake (AI) for adult men (550 mg choline/day) (20). Participants were provided with all foods, drinks, and snacks for the duration of each of the three 2-week controlled feeding arms followed by a 2-week washout period in which the participants returned to their usual diet. These diets were identical except for the modification of the amount of choline (as choline chloride), added to three bread rolls per day. Choline content and stability during choline storage in the bread rolls were determined separately for each batch produced. The amount of choline in each diet was not adjusted for subject sex or BMI. All study diets were designed to provide approximately equal amounts of betaine (average of 147 mg/day betaine. (See **Supplementary Table 1** for the nutrient breakdown of diets, including choline and betaine, and **Supplementary Table 2** for nutrient breakdown and ingredients of bread rolls). Study participants received, in random order, each of the 3 dietary arms, delivering: 5.1 mmol choline/day (531 mg, 97% AI), 2.5 mmol choline/day (47% AI), or 1.3 mmol choline/day (25% AI). There was a minimum of 2 weeks of washout between arms during which the participants returned to their usual diet. On Day 12 of each dietary arm, in addition to the food supplied, subjects consumed a single bolus of 2.2 mmol trimethyl-d_9_-choline (as 98% D, methyl-perdeuterated chloride; Cambridge Isotope Laboratories; Tewksbury, MA, USA). This dose is based on previous reports (21), with an additional ∼25% to account for differential absorption between intravenous and oral delivery. We confirmed total choline and betaine content of diets using duplicate food portions analyzed by liquid chromatography-stable isotope dilution-multiple reaction monitoring mass spectrometry (LC-SID-MRM/MS) as previously described (22). Riboflavin (30 mg/day) was added to the bread rolls and was used as a compliance biomarker via 24-hour urine collection measures (23–25). 24h-urinary excretion of riboflavin was measured at baseline (Day 1) and on Day 15 of each arm. Compliance was assessed based on an increase in riboflavin excretion of ≥ 1 mg/mL over individual baselines (24). Participants who were non-compliant in two arms were excluded from all the analysis. Although all participants completed the 3 study arms, compliance was assessed only after study completion based on riboflavin measurements. Three participants were excluded due to non-compliance (1 man, 1 premenopausal woman, and 1 menopausal woman).

Blood samples were collected by venipuncture on days 1, 12, 13, and 15.

The protocol was approved by the Institutional Review Board at the University of North Carolina at Chapel Hill and informed consent was obtained.

### Subject recruitment

#### Inclusion/exclusion criteria

All participants were recruited from the greater Kannapolis-Charlotte, NC metro area and screened by a physician, and by self-report assessment. Clinical laboratory tests were run, including a comprehensive metabolic panel.

Inclusion criteria were the age range of 20-67 years and BMI range of 20-30 kg/m^2^. Exclusion criteria included medication known to alter or damage liver or muscle health, and to have the potential to alter choline metabolism. Participants with chronic systemic disease, a history of hepatic or renal disease, being a substance abuser, current smoker, or alcohol consumption of more than 2 alcoholic beverages per day (14 per week) were also excluded, as were participants consuming choline-containing dietary supplements, frequently consuming a diet that might interfere with the study, or having allergies to soy proteins or any foods in the required diet. Subjects who performed intense exercise of more than 1 hour every day or other intense muscle building exercises (such as weightlifting low weight maintenance repetitions) were also excluded.

Women who were pregnant, breastfeeding, or planning to become pregnant were excluded due to the potential risk to the fetus/baby of a low choline diet. Ultimately, 101 healthy volunteers (50 premenopausal women, 20 postmenopausal women, 1 perimenopausal woman, 30 men) were enrolled (**Table 1**). A CONSORT diagram is included (**Figure 1**). Since participant recruitment occurred during the COVID-19 pandemic, study visits were suspended beginning in March 2022 and resumed in June 2023. COVID-19 symptoms and travel screenings were conducted within 24 hours prior to each visit and again upon arrival. Body temperature was checked at entry, and masks were required for both staff and participants throughout the visit, with additional eye protection used as needed. Social distancing was maintained by modifying procedures—for example, participants self-operated the body composition scale and stadiometer under staff supervision. All surfaces and equipment were sanitized before and after each visit. Two IRB-approved protocol amendments were submitted to implement and formalize these safety measures.

**Table 1.** Participant characteristics. Follicle stimulating hormone (FSH) levels as measured by LabCorp were used to classify menopausal status.

|  | **Men** | **Premenopausal women** | **Menopausal women** | **Perimenopausal women** |
| --- | --- | --- | --- | --- |
| **Number of participants (n)** | 24 | 39 | 14 | 1 |
| **Age (years)** | 36 ± 9.7 | 35 ± 8.5 | 53 ± 6.8 | 49 |
| **Height (centimeters)** | 177 ± 6 | 165 ± 5 | 166 ± 5.8 | 161 |
| **Weight (kilograms)** | 77 ± 9 | 70 ± 6.9 | 68 ± 7.8 | 74.20 |
| **BMI kg/m<sup>2</sup>** | 24 ± 2 | 26 ± 2.8 | 25 ± 3 | 28.60 |
| **Ethnicity (white/black/hispanic)** | 21/2/1 | 26/7/6 | 11/3/0 | 1/0/0 |
| **FSH (mIU/mL)** | N/A | 6 ± 3.30 | 79. ± 31.4 | 23 |
| **Estrogen (mIU/mL)** | N/A | 212 ± 150.6 | 96.79 ± 66.5 | 128 |

**Figure 1.**
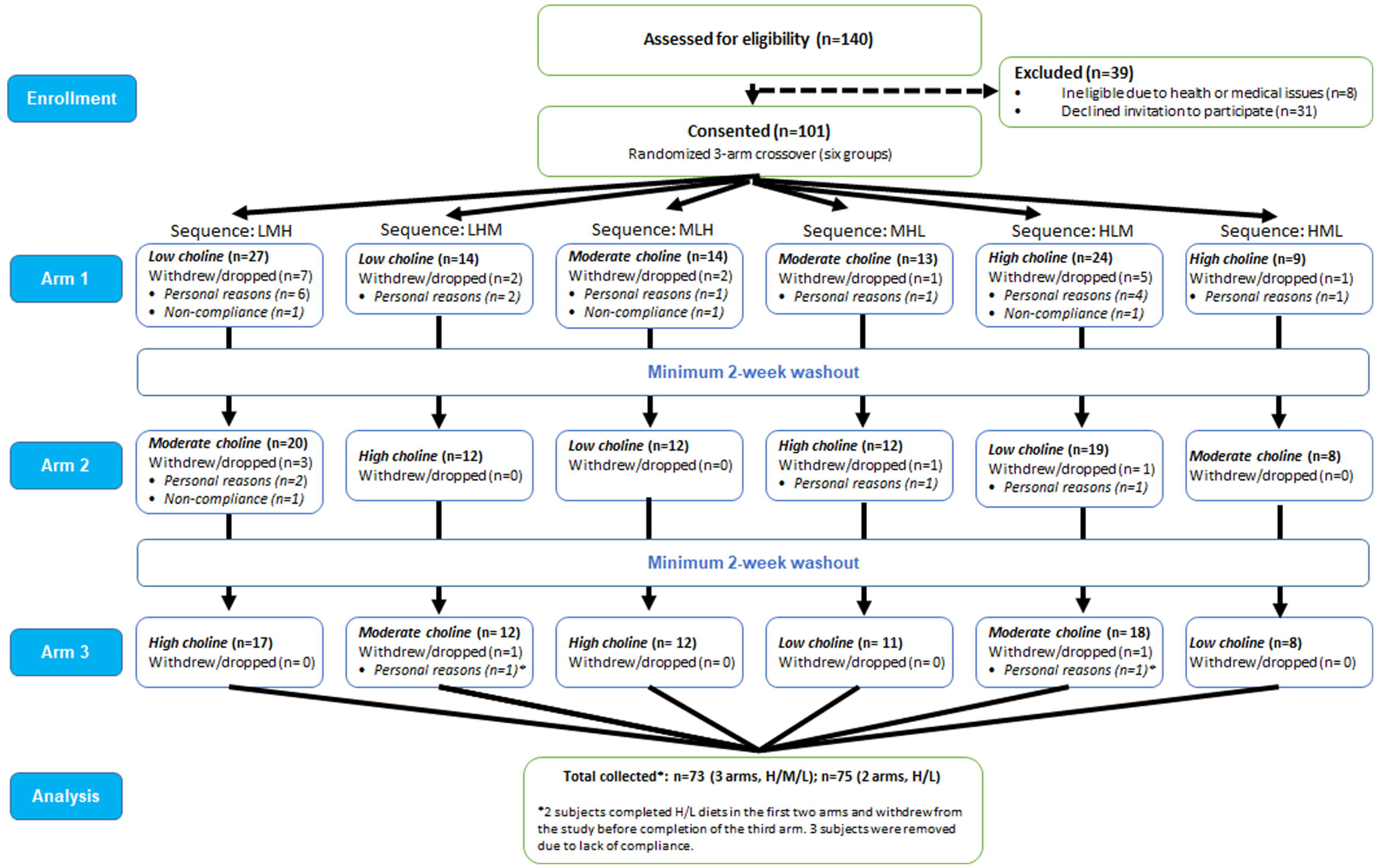
CONSORT diagram. Flowchart of participants through the study. L: 25%AI choline intake; M: 50% AI choline intake; H: 100% AI choline intake

#### Sample collection

All subjects were seen at the Human Research Core at the University of North Carolina Chapel Hill, Nutrition Research Institute, Kannapolis, NC and fasted overnight prior to blood collection. Whole blood was collected in serum-separating tubes (SST) for hormone measurements and lithium heparin tubes for plasma metabolite analysis. Tubes were gently inverted to mix. Lithium heparin tubes were immediately put on wet ice and centrifuged at 2200 × g at 4 °C for 10 minutes, then returned to the ice bath. Plasma samples were pooled, aliquoted and frozen at - 80 °C within an hour of collection time. SST tubes were allowed to clot at room temperature then centrifuged at 2200 × g at 4 °C for 10 minutes. SST tubes were sent for a comprehensive metabolic panel and creatine kinase testing to the clinical laboratories at Lab Corporation of America, Burlington, NC, which is both Clinical Laboratory Improvement Act (CLIA ID 34D0655059) and College of American Pathologists (CAP No. 1396901) accredited. At enrollment, follicle-stimulating hormone (FSH) and estrogen were also measured in female participants.

Urine was collected by participants into a commode specimen collector and immediately poured into a 3-L plastic jug that contained 3 mL acetic acid. The plastic jug was kept cool with ice packs in an insulated cooler during the 24-h collection. On completion, the total volume and collection times were noted. Aliquots of 1 mL were created with and without formic acid (10 µL) and stored at -80 °C.

Tanita® Body Composition Analyzer was used for weight and body composition measures following manufacturer instructions. Participants were weighed fully clothed but with shoes and socks removed. Height used for entry in analyzer was measured with shoes off and using a wall-mounted stadiometer.

The Automated Self-Administered 24-hr (ASA24®) Dietary Assessment was completed by participants prior to enrolling in the study. They entered 3 days of dietary recall. The first day was entered during the screening visit so that participants would have direct instruction and guidance on how to enter the data. They were instructed to complete two additional “typical” meal days prior to start of study.

### FibroScan***®*** measurements

Liver stiffness was assessed by transient elastography (FibroScan®, Echosens, Paris). The examination was performed at the Human Research Core at the University of North Carolina Chapel Hill, Nutrition Research Institute, Kannapolis, NC, by technicians or physicians blinded to the participant’s choline intake status. The FibroScan used was a FibroScan® 502 Touch model, equipped with S, M, and XL probes. Probes were automatically selected by the software’s tool. The examination procedure was as follows: participants were asked to fast one night before the examination, then placed in a supine position with their right arm fully abducted. Participants were asked to hold their breath for 10-15 seconds, and at least 12 measurements per participant were collected on Day 1 and Day 15 of each arm. Measurements were performed by scanning the liver through an intercostal space. We repeated the procedure and recorded at least four sets of 12 measurements per collection day. The raw ultrasonic radio frequency signals were stored in the Fibroscan file. The controlled attenuation parameter (CAP) was computed only when the measurement was valid. The final CAP results were expressed in dB/m and corresponded to the average of valid individual measurements. Importantly, in our study, Fibroscan was not used to diagnose steatosis (26, 27), but rather to detect significant changes in liver fat content resulting from the choline dietary intervention.

#### Mass spectrometry for plasma choline metabolites and urine riboflavin

Samples were prepared by spiking with stable isotope-labeled internal standards, followed by extraction using a protocol we established (28, 29). Briefly, each sample was combined with four volumes of methanol:chloroform (2:1, v/v), vortexed, and incubated at −20 °C for 2 to 24 hours before a second vortexing step. The resulting supernatant was collected, and the remaining pellet was subjected to a secondary extraction using methanol:chloroform:water (2:1:0.8, v/v/v). Supernatants from both steps were pooled and subjected to phase separation by the addition of water and chloroform. The aqueous layer, containing choline and betaine, was transferred to HPLC vials for analysis. The organic phase, containing phosphatidylcholine and sphingomyelin, was diluted in acetonitrile and similarly transferred to HPLC vials. Calibration standards containing known analyte concentration and internal standards were prepared in parallel and processed using the same extraction steps.

Quantification of analytes was performed by liquid chromatography coupled with stable isotope dilution tandem multiple reaction monitoring mass spectrometry (LC-SID-MRM/MS) for both aqueous and organic fractions, as previously established (22, 28, 29). Chromatographic separation was achieved using and Acquity HILIC column (1.7 µm, 2.1 × 50 mm; Waters Corp.) on a Waters ACQUITY UPLC system, operated at a flow rate of 0.37 mL/min and maintained at 40 °C. For aqueous-phase analysis, the mobile phases consisted of A) 100% water containing 0.125% formic acid, and B) 90:10 acetonitrile:water with 10 mM ammonium formate and 0.125% formic acid. Mobile phase A for organic analytes was 10% acetonitrile/90% water and 0.125% formic acid and B was the same as for aqueous metabolites. As described previously (28), phosphatidylcholine undergoes in-source fragmentation and is detected in the form of phosphocholine. Metabolites and their corresponding isotope-labeled standards were monitored on a Waters triple quadrupole mass spectrometer, using specific precursor-product ion transitions. Analyte concentrations were calculated from the ratio of the signal intensity of each analyte to its internal standard, referenced against aa standard curve generated from known concentrations. The isotopic enrichment ratio (IER) was determined by calculating the proportion of labeled metabolite relative to the total (labeled plus unlabeled) pool, providing a measure of precursor availability within the metabolic pathway (29, 30).

#### Mass spectrometry for homocysteine

Total plasma homocysteine (tHcy), representing the sum of all reduced and oxidized forms following disulfide bond cleavage, was quantified using LC-ESI-MS/MS, as previously described with some minor modification (31). Plasma (10 µL) was prepared by incubation on ice for 20 minutes with a aqueous solution (120 µL) containing internal standard d8-homocystine (Cambridge isotopes) and dithiothreitol (DTT). The mixture was then deproteinized by the addition of acetonitrile (400 µL) that was mixed and then centrifuged at 14,000 rpm for 10 minutes at 4°C. The resulting clear extract was transferred to a 96 well plate and loaded into a refrigerated autosampler for analysis by the mass spectrometry system (4000 QTRAP, AB Sciex LLC, Framingham, MA). Data was acquired and processed using Sciex Analyst v1.6.24 (AB Sciex LLC, Framingham, MA).

#### Receiver operating characteristic (ROC) data analysis

Data preprocessing was completed using MetIDQ Carbon software for peak extraction, automatic checking of peak alignment, normalization, and background deduction of mass spectrometry raw data. Receiver operating characteristics (ROC) curves were generated using MetaboAnalyst (32). All data was uploaded to the web server followed by a data integrity check with missing value imputations. All data was normalized using log transformation followed by auto scaling. Multivariate ROC curves were generated using a linear SVM algorithm and predicted class probabilities were determined using 100 cross-validations. The area under the curve (AUC) was calculated to assess model discrimination. To guard against overfitting, model complexity was optimized to minimize validation errors, and permutation testing (n = 1000) was performed to confirm that the observed AUC was not due to chance. Results are reported as mean AUC values with confidence intervals based on cross-validation replicates.

#### Statistical analysis

We conducted a repeated-measures mixed-effects analysis with Geisser-Greenhouse correction using GraphPad Prism 10 (GraphPad Software Inc., California, USA). This model is well-suited for our randomized design and accommodates random missing values without the need for imputation, making it preferable to traditional ANOVA context. For multiple comparisons, we used Tukey’s test.

Linear mixed effects modelling was performed in R (version 4.3.0) using lme4 (version 1.1-35.3) and lmerTest (version 3.1-3) packages(33). Plasma concentrations of choline and choline-related metabolites were modeled as the continuous outcome variables. Fixed effects included choline intake (modeled as three levels: 100% AI, 50% AI, and 25% AI) and sex/hormonal status, represented as a four-factor categorical variable (male, female-premenopausal, female-perimenopausal, and female-postmenopausal). To account for the correlation between repeated measures or multiple observations from the same individual, a random intercept was included for participant ID, capturing individual-level variation in baseline metabolite concentrations. P-values for fixed effects were obtained using Satterthwaite’s approximation for degrees of freedom, as implemented in the lmerTest package. All models were fit using restricted maximum likelihood (REML) estimation (34). P values below 0.05 were considered significant.

## RESULTS

Plasma choline concentrations are tightly regulated and may not reliably reflect dietary choline intake on their own (17). To address this limitation, we evaluated whether the combined measurement of plasma choline, betaine and PtdCho concentrations could more accurately reflect dietary choline intake.

### Isotopic enrichment ratios (IER) of betaine respond as a function of the dietary choline intake after a d_9_-choline bolus

Plasma concentration of choline, betaine and PtdCho concentrations were measured by MS at base line (pre-bolus; d_0_) and 24-hours post-bolus (d_9_) following administration of d_9_-choline bolus. Isotopic enrichment ratios (IER) were calculated as d_9_-metabolite/(d_0_+ d_9_-metabolite), reflecting the proportion of labeled compound in the circulating pool. Metabolite concentrations are presented in **Table 2**. Due to the absence of detectable d_9_-choline in plasma at 24 hours, these data are included in **Supplementary Table 3**.

**Table 2.** Concentration of plasma betaine and PtdCho measured by mass spectrometry. Betaine and PtdCho were measured by MS at basal and 24 hours post bolus (2.2 mmol d_9_-choline). Values are mean ± SEM. Data for the single perimenopausal female are include in All but not listed separately. Men n=23, Menopausal women n=13, premenopausal women n=38 and perimenopausal woman n=1.

BETAINE AND PtdCho PLASMA CONCENTRATIONS AFTER d<sub>9</sub>-choline BOLUS
|  | CHOLINE DIET 25% | CHOLINE DIET 50% | CHOLINE DIET 100% |
| --- | --- | --- | --- |
| **d<sub>0</sub>-betaine (μm)** |  |  |  |
| **MEN** | 63.5 ± 10.3 | 74 ± 13 | 92.5 ± 18 |
| **PREMENOPAUSAL (F)** | 45 ± 15 | 52 ± 18 | 68 ± 25 |
| **MENOPAUSAL (F)** | 53.4 ± 12 | 61.8 ± 15 | 80.5 ± 23 |
| **ALL** | 52 ± 15 | 61 ± 18 | 78 ± 25 |
| **d<sub>9</sub>-betaine (μm)** |  |  |  |
| **MEN** | 1.6 ± 0.47 | 1.9 ± 1.20 | 2.0 ± .77 |
| **PREMENOPAUSAL (W)** | 1.18 ± 0.68 | 1.33 ± 0.67 | 1.63 ± 0.80 |
| **MENOPAUSAL** | 1.26 ± 0.66 | 1.44 ± 0.71 | 1.73 ± 0.80 |
| **ALL** | 1.3 ± 0.64 | 1.6 ± 0.92 | 1.8 ± 0.81 |
| **IER-betaine (μm)** |  |  |  |
| **MEN** | 0.024 ± 0.004 | 0.025 ± 0.010 | 0.021 ± 0.006 |
| **PREMENOPAUSAL (W)** | 0.023 ± 0.68 | 0.024 ± 0.006 | 0.022 ± 0.005 |
| **MENOPAUSAL (W)** | 0.022 ± 0.008 | 0.021 ± 0.010 | 0.020 ± 0.005 |
| **ALL** | 0.022 ± 0.008 | 0.024 ± 0.008 | 0.002 ± 0.005 |
| **d<sub>0</sub>-PtdCho (μm)** |  |  |  |
| **MEN** | 2055 ± 408 | 2132 ± 481 | 2056 ± 424 |
| **PREMENOPAUSAL (W)** | 2172 ± 474 | 2240 ± 526 | 2208 ± 551 |
| **MENOPAUSAL (W)** | 2583 ± 421 | 2636 ± 453 | 2707 ± 607 |
| **ALL** | 2209 ± 476 | 2277 ± 524 | 2249 ± 565 |
| **d<sub>9</sub>- PtdCho (μm)** |  |  |  |
| **MEN** | 27.1 ± 7.02 | 26.8 ± 9.74 | 21.8 ± 7.94 |
| **PREMENOPAUSAL (W)** | 24.3 ± 8.36 | 25 ± 7.92 | 22.6 ± 5.88 |
| **MENOPAUSAL (W)** | 25.9 ± 6.82 | 26.4 ± 10.6 | 25 ± 3.85 |
| **ALL** | 26 ± 7.7 | 26 ± 8.9 | 23 ± 6.4 |
| **IER- PtdCho (μm)** |  |  |  |
| **MEN** | 0.013 ± 0.002 | 0.012 ± 0.003 | 0.010 ± 0.002 |
| **PREMENOPAUSAL (W)** | 0.011 ± 0.003 | 0.011 ± 0.002 | 0.010 ± 0.002 |
| **MENOPAUSAL (W)** | 0.010 ± 0.003 | 0.010 ± 0.003 | 0.009 ± 0.001 |
| **ALL** | 0.012 ± 0.003 | 0.011 ± 0.003 | 0.010 ± 0.002 |

A mixed-effect analysis adjusted for false discovery rate revealed a significant diet-dependent effect on baseline d_0_-choline and betaine concentrations (**q<0.0001**) but not for PtdCho. Pairwise comparisons for PtdCho were non-significant across dietary groups: 25% AI vs 50% AI (q=0.277), 25% AI vs 100% AI (q=0.531), and 50% AI vs 100% AI (q=0.533) (**Supplementary Table 3 and Table 2**). These findings indicate that baseline plasma concentrations of choline and betaine, but not PtdCho, reflect dietary choline intake.

For the labeled metabolites, d_9_-betaine showed a strong intake-dependent response (overall **p<0.0001**), with significant pairwise differences between 25% AI and both 50% AI (**q=0.0041**) and 100% AI (**q=<0.0001**), as well as between 50% AI and 100% AI (**q=0.0033**).

d_9_-PtdCho also varied by intake level (**p=0.0017**), with significant differences observed between 25% AI vs 100% AI (**q=0.001**), and 50% AI 100% AI (**q=0.0006**), but not between 25% AI vs 50% AI (q=0.2657).

The values for IER-betaine demonstrated a significant overall effect of diet (**p=0.026**), with differences between 25% AI vs 50% and 100% AI (**both q<0.01**), but not between 25% AI vs 50% AI.

IER-PtdCho showed a significant overall effect on the mixed-effects model (**p=0.0001**). Pairwise comparisons indicated highly significant differences between 25% AI vs 100% AI and 50% AI vs 100% AI, we found highly significant differences (**q=0.0001** and **q=0.0005**, respectively) and not significant differences between 25% AI vs 50% AI (q=0.126).

Together, these findings confirm that d_9_-betaine and d_9_-PtdCho, and their respective IERs, are responsive to dietary choline intake, particularly in comparisons between the 25% AI and 100%

AI groups. These results aligned with previous pilot findings (29), now supported by increased number of participants. Since the above analyses reflect the response to a single d_9_-choline bolus administered on Day 12 and evaluated at Day 13, we next assessed whether steady-state plasma concentrations of choline, betaine, PtdCho and total Hcy (tHcy) at the end of each arm (Day 15) also reflected dietary intake by using targeted metabolomics.

### Plasma choline and betaine concentrations reflect dietary choline intake

Using targeted MS, we assessed plasma concentrations of choline, betaine, PtdCho and tHcy on Day 15, marking the end of each study arm. Because choline metabolism differs between men, premenopausal women and menopausal women based on sex and hormonal status (17), our evaluations consider these factors.

We observed that plasma choline concentrations were statistically significant across dietary choline intake levels (25%, 50%, and 100% AI) in men, with the exception of the comparison between 25% and 50% AI (**Fig. 2A).** In premenopausal women, significant differences were also detected across the choline intake levels (**Fig. 2B**). However, in menopausal women, a significant difference was only observed between the 25% AI and 100% AI groups (**Fig. 2C).** When data from all participants were combined, plasma choline concentrations differed significantly and in the same direction as the dietary choline intake (**Fig. 2D).**

**Figure 2.**
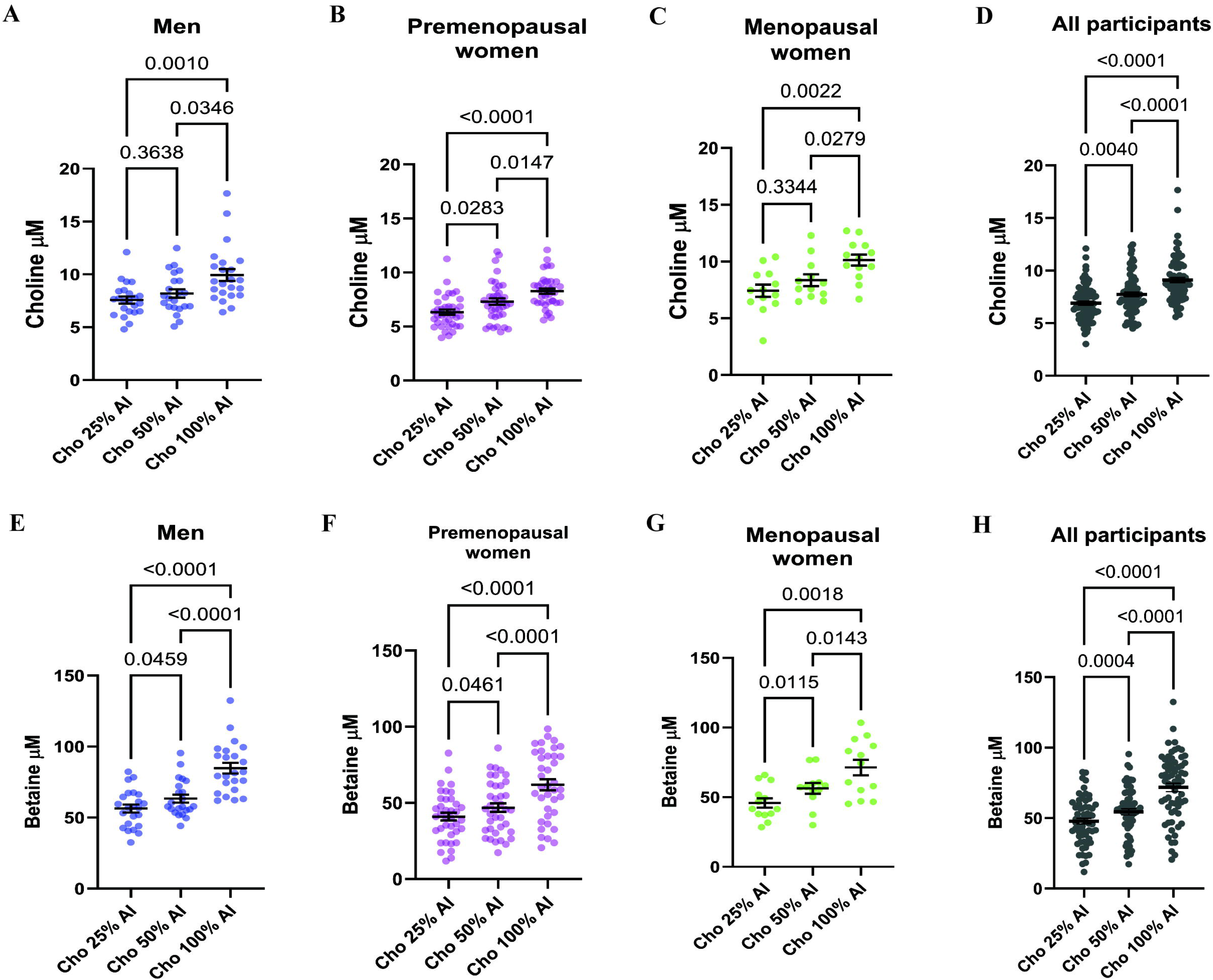
Plasma choline and betaine concentrations by sex and dietary choline intake level. Plasma choline concentrations (**A-D**) and plasma betaine concentration (**E-H**) are shown at the end of each dietary arm (25%, 50% and 100% of the Adequate Intake (AI) for choline). Graphs show data stratified by sex and menopausal status, men, premenopausal women, menopausal women and all participants. Each dot represents one participant; bar represents the group mean ± SEM. Data was analyzed using a linear mixed-effects model with fixed effect for diet and subject as a random effect with Geisser-Greenhouse correction. Post hoc comparisons were performed using Tukey’s test and reported p values are adjusted for multiple comparisons. Sample size: men n= 23, premenopausal women n=38 and menopausal women n=13. Note that for the Arm Cho 50% AI, plasma samples could not be obtained for one man and one menopausal woman. Perimenopausal woman included in “all participants

As described in the methods, all study diets were designed to provide approximately equal amounts of betaine (∼147 mg betaine per day). Therefore, observed changes in plasma betaine concentrations are a response to differences in dietary choline intake. By Day 15, plasma betaine levels varied significantly across the intake levels in all the groups, except for men comparing 25% AI to 50% AI (**Figure 2E**). In both premenopausal and menopausal women, betaine concentrations differed significantly by dietary intake level (**Fig.2 F-G).** When data were analyzed across all participants, significant differences in plasma betaine were also observed in response to dietary choline intake (**Fig. 2H).**

In contrast, plasma PtdCho concentrations at Day 15 did not differ significantly across choline intake levels when stratified by sex, hormonal status, or in the overall cohort (**Supplementary Fig. 1A-D**). Given that both choline and betaine are inversely associated with plasma total homocysteine (tHcy) concentrations (35, 36), we measured tHcy at Day 15 of each dietary arm (**Supplementary Fig1. E-H**). A statistically significant reduction in tHcy was observed when comparing the 25% AI vs 50% AI groups (**p=0.033**) in the combined all participants analysis; however, no significant differences were detected in other comparisons. Collectively, these findings support that plasma choline and betaine concentrations are responsive to dietary choline intake across the full cohort and within subgroups defined by sex and hormonal status.

### Fibroscan™ predicts choline dietary intake using changes in liver stiffness in a subset of participants

We then asked whether additional clinical markers could enhance the prediction of dietary choline intake. We previously demonstrated that severe dietary choline restriction, specifically 10% of the AI, was associated with the development of steatosis and liver injury (17, 37). In the current study, the lowest choline intake was 25% of AI. Given this, we investigated whether this level of choline restriction was sufficient to elicit early markers of liver dysfunction. To assess liver function, we measured the circulating concentration of AST and ALT at the end of each arm. No statistically significant differences were observed among the three dietary choline groups (**Supplementary Fig 2A-H**). These findings suggest that choline intakes above 25% of the AI, when consumed over a two-week period, were sufficient to maintain liver health and prevent elevation in liver enzymes. This contrasts with our prior study in which prolonged consumption, up to 7 weeks, of diets providing only 10% of the AI led to clinically evident liver injury (17, 37).

Since we did not observe abnormal liver enzymes in circulating plasma, we tested whether Fibroscan, a non-invasive approach to assess liver fat (38), could detect liver fat changes in response to the different choline dietary intakes. We measured liver fat as controlled attenuation parameter (CAP) as a reflection of liver fat. We calculated the final value for each arm by averaging four independent sets, each consisting of 12 measurements. First, we analyzed whether participants exhibited significant differences on Day 1 of each arm, with the objective of correcting liver fat differences at the start of the study. We analyzed results stratified by sex and hormonal status, and we did not find differences on Day 1 when comparing CAP values between 25% AI *vs* 50% AI *vs* 100% AI (**Supplementary Figure 3A-C**). However, when we pooled all the participants, we found significant differences on Day 1 when starting the 50% AI compared to 25% AI arm (**p=0.0124**; **Supplementary Figure 3D**). These findings showed us that the CAP values, reflecting liver fat, on Day 1 were influenced by their regular dietary intake prior to the study enrollment. As a result, we focused our analysis on Day 15 CAP measurements, after participants had consumed controlled diets for 15 consecutive days.

When analyzing the impact of consuming 50% AI choline diet, we found a subset of participants that showed hepatic fat accumulation, while others remained unaffected (**Supplementary Figure 3E**). These interindividual differences in response to choline are likely to be multifactorial but at least in part driven by genetic variants influencing choline metabolism (39, 40).

Given our previous results, we asked if there was an increase in liver fat accumulation from 100% AI compared to 25% AI arms. Participants were classified as responders if they exhibited a ≥ 10% increase in CAP values between the 100% AI arm vs 25% AI. For this analysis, we used participants with complete Fibroscan assessments. Among 72 participants, 30 individuals (**41.6%**), exhibited an increase of at least 10% in liver fat when consuming a 25% AI diet **(Figure 3)**; this group was classified as “responders” and the rest of the participants did not meet this criterion (**Supplementary Figure 3F**).

**Figure 3.**
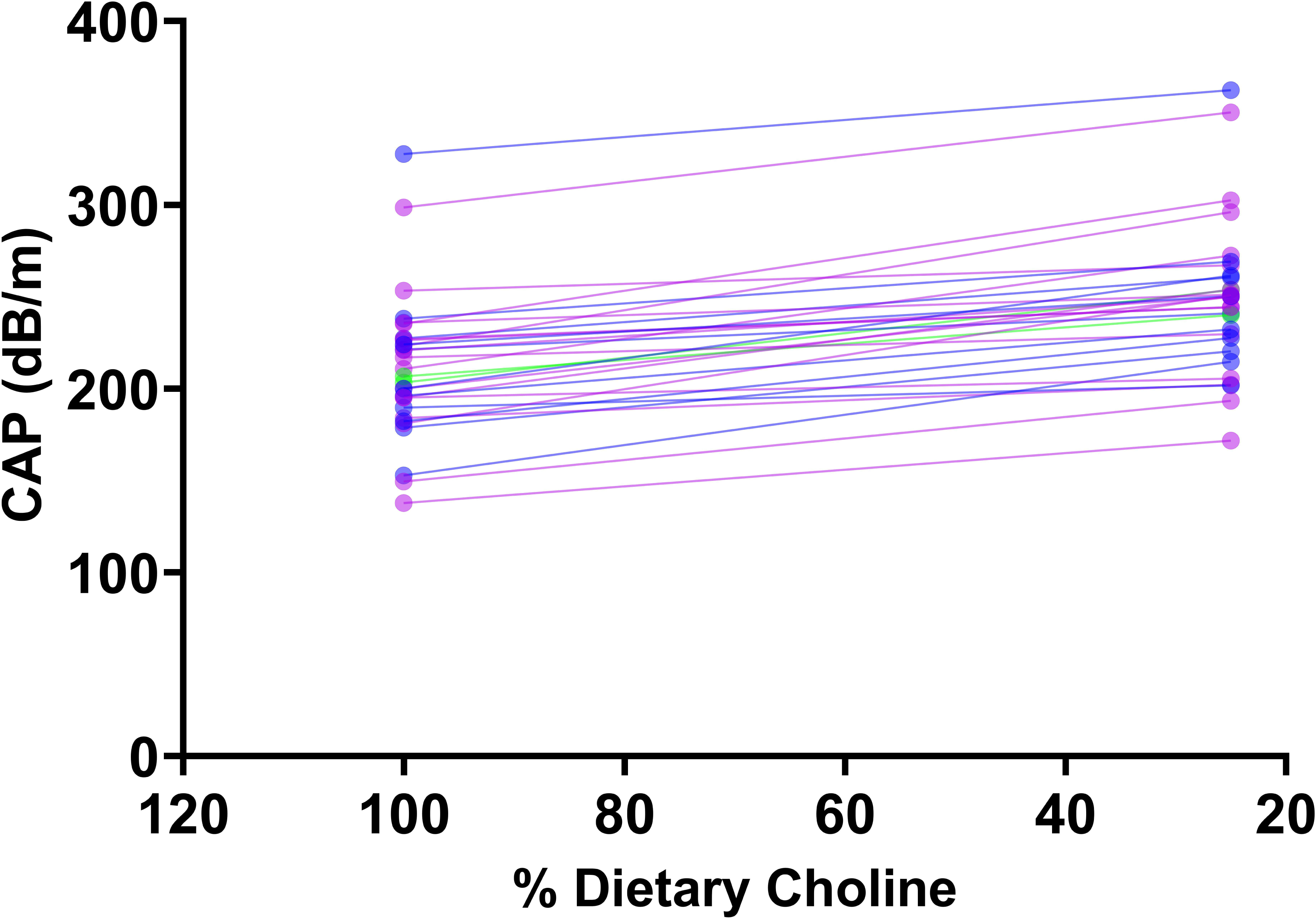
Individual slope plots showing changes in controlled attenuation parameter (CAP) measured by Fibroscan at the end of the 100% AI and 25% AI dietary choline intake. Each line represents one participant, with CAP values (dB/m) measured at the end of each condition. These participants showed at least 10% increase in liver fat when consuming 25% AI compared with 100% AI.

Altogether, these findings indicate that liver fat serves as a marker of dietary choline intake in a specific subset of participants who demonstrate an increase in CAP averages when consuming 25% AI choline. However, the utility of Fibroscan as a standalone tool for assessing dietary choline intake is limited, given that a considerable portion of participants exhibited no or minimal changes in liver fat across choline dietary intake.

### Association between dietary choline intake and liver fat content in a linear mixed-effects model

Since we had plasma choline metabolites and Fibroscan CAP numbers at the end of each arm, we conducted linear regression analysis to assess the effect of dietary choline intake (25% AI and 50% AI) compared to the 100% AI reference group on plasma metabolites (choline, betaine, PtdCho and tHcy) and CAP values as a reflection of liver fat. Both plasma and betaine concentrations were significantly lower in 25% AI and 50% AI groups relative to 100% AI. In contrast, PtdCho concentrations did not differ significantly by dietary choline intake level (**Table 3**).

**Table 3.** Linear mixed-effects model estimates (β coefficients and 95% confidence intervals) comparing circulating metabolite concentrations of choline, betaine and PtdCho across dietary choline intake levels. The 100% Adequate Intake (AI) condition was used as the reference group. Fixed effects included choline intake and sex/hormonal status, while participant ID was included as a random intercept to account for repeated measures. Plasma choline and betaine concentrations were significantly lower in the 25% and 50% AI groups compared with 100% AI.

| Metabolite |  | Beta(LCL,UCL) | p |
| --- | --- | --- | --- |
| Choline | ref: 100%AI |  |  |
|  | 25% AI | -2.20 (-2.72,-1.68) | 0.00000000000000544313 |
|  | 50%AI | -1.35 (-1.87,-0.83) | 0.00000120299574607583 |
| Betaine | ref: 100%AI |  |  |
|  | 25% AI | -24.05 (-27.72,-20.39) | 0.0000000000000000000000011 |
|  | 50%AI | -16.82 (-20.52,-13.12) | 0.000000000000000201426475108 |
| Ptd Choline | ref: 100%AI |  |  |
|  | 25% AI | -38.22 (-255.79,179.09) | 0.7308934 |
|  | 50%AI | 37.23 (-181.83,256.32) | 0.73968961 |

Total homocysteine was modestly elevated in the 25% AI group which is consistent with previous reports where total choline and betaine intake was inversely associated with tHcy (35). No statistically significant differences in liver fat, as measured by Fibroscan CAP values at Day 15, were observed across the groups. Similarly, a change in CAP (ΔCAP) did not differ significantly between groups (**Supplementary Table 4**).

These results indicate that plasma choline and betaine concentrations are sensitive to moderate changes in dietary choline intake.

### Use of plasma choline and betaine concentration shows high specificity in predicting dietary choline intake

Across multiple analytical approaches shown in this study, our findings consistently demonstrated that plasma choline and betaine concentrations responded to dietary choline intake, with the most pronounced differences observed between the lowest (25% AI) and highest (100% AI) choline intake levels. To further evaluate the utility of these metabolites in classifying dietary choline intake, we performed Receiver Operator Characteristics (ROC) curve analysis using plasma values from participants completing the 25% AI and 100% AI arms.

Plasma choline alone yielded an area under the curve (AUC) of 0.811 (95% CI: 0.74-0.87) indicating a good discriminatory capacity (**Figure 4A**). Similarly, plasma betaine exhibited an AUC of 0.807 (95% CI 0.72-0.87) (**Figure 4B**), supporting its role as an individual predictor. When choline and betaine were combined the AUC increased to 0.83 (95% CU 0.73-0.91), indicating excellent predictive accuracy with a confidence interval spanning from acceptable to outstanding discrimination (**Figure 4C**).

**Figure 4.**
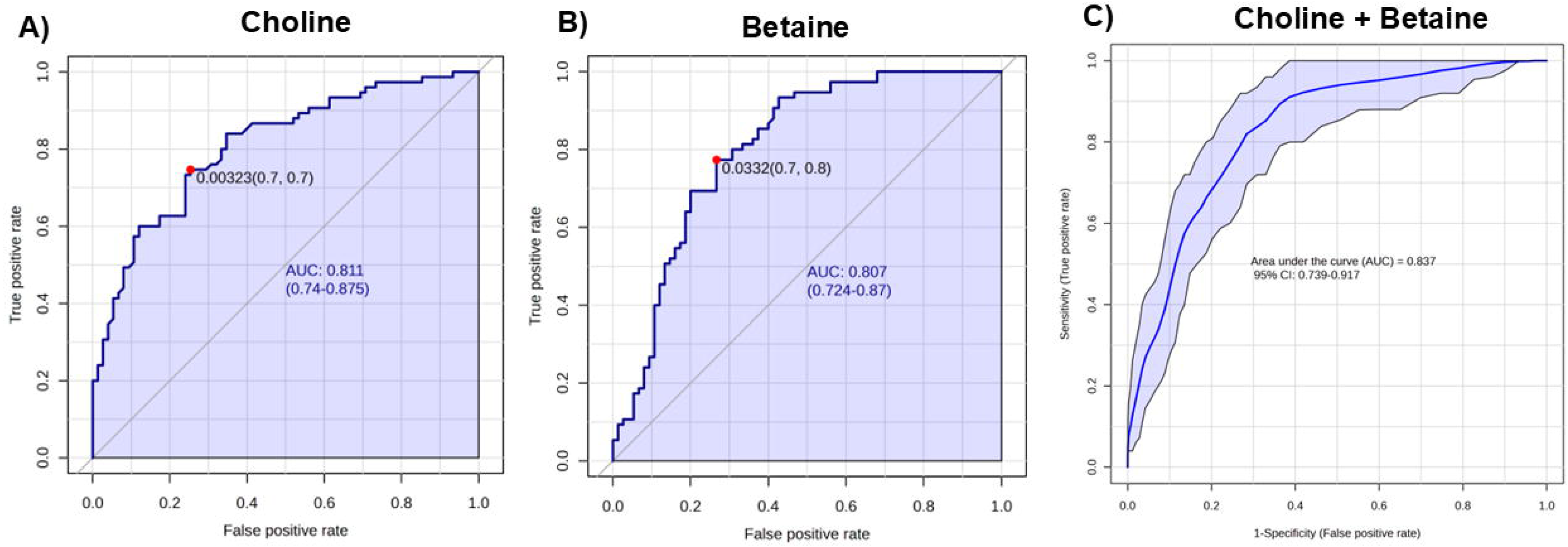
Receiver operating characteristic (ROC) curve assessing the performance of choline, betaine and choline + betaine in reflecting dietary choline intake. **A)** ROC curve for plasma choline as a predictor of dietary choline intake. The area under the curve (AUC) was 0.81 (95% CI: 0.735, 0.872), indicating a good discriminatory capacity. The optimal cutoff (red dot) corresponds to a false positive rate of 0.32% and a balances sensitivity and specificity values of 70% each. The shaded blue region represents the 95% confidence band for the ROC curve. **B)** ROC curve for plasma betaine as a predictor of dietary choline intake. The AUC was 0.803 (95% CI: 0.724, 0.873), indicating good discriminatory ability. The optimal threshold (red dot) was identified at a false positive rate at 3.3% corresponding to sensitivity of 70% and specificity to 80%, and it is determined by maximizing Youden’s J (using MetaboAnalyst). **C)** ROC curve evaluating the performance of choline and betaine in reflecting dietary choline intake. The AUC was 0.837 (95% CI:0.739, 0.917), indicating excellent discrimination. The shaded blue denotes the 95% confidence band.

These findings suggest that choline and betaine, alone and in combination, are robust markers of dietary choline intake. We acknowledge that a limitation of our approach is the lack of independent manipulation of dietary betaine, which may contribute to collinearity in the predictive model. Nonetheless, the consistency and strength of the ROC performance supports the value of these markers in assessing choline dietary intake under controlled conditions.

## DISCUSSION

The role of choline in human nutrition is well-established, particularly in relation to brain and cognitive function, liver metabolism, and muscle health (2, 7, 18, 41, 42). Despite this, accurate determination of dietary choline intake remains a methodological challenge. Early studies demonstrated that consuming only 10% of the AI of choline for six weeks can result in the development of hepatic steatosis and liver injury (17). However, the effects of a moderate reduction in choline intake over shorter duration may provide a more realistic insight into habitual dietary choline intake patterns. With this perspective in mind, the present study employed a combination of approaches to identify early and sensitive indicators of low dietary choline intake, that could support more refined dietary guidance.

In our first approach, we administer a single bolus of deuterium-labeled choline (d_9_-choline chloride) as a tracer to evaluate whether plasma levels of d_9_-choline, d_9_-betaine or d_9_-PtdCho, measured 24 hours post-dose, reflected dietary choline intake. While d_9_-betaine or d_9_-PtdCho were reliably detected and demonstrated significant differences, d_9_-choline was not detectable in most participants at the 24-hours post bolus (the liver rapidly clears choline from blood after oral administration (43). These findings are consistent with our earlier interim analysis during method development where 6 hours post bolus showed modest changes and the values were low and showed minimal discrimination by intake level (29).

d_9_-PtdCho concentrations in plasma were comparatively high, though they showed weaker discrimination across intake levels. This metabolic partitioning is consistent with known kinetics of choline utilization, at lower concentrations of choline, the liver phosphorylates it and produces phosphatidylcholine; at higher concentrations of choline, choline kinase becomes saturated and most choline is oxidized to form betaine (43). Notably, as outlined in the Methods section, we calculate the same amount of betaine intake in the diets provided, enhancing our confidence in attributing changes to dietary choline intake.

Previously, tracer experiments in men who consumed d_9_-choline revealed that this method can discern choline metabolism variations driven by genetic variants in the MTHFR (677CC or 677TT) gene (44). One of the major contributions of our approach it is the potential for reflecting dietary choline intake with a single bolus of d_9_-choline chloride, irrespective of sex or genotype. Consolidating these findings highlights the necessity of evaluating established genetic variants linked to increasing the requirements (45, 46). However, the cost of performing tracer experiments limits its use as a reflector of recent dietary choline intake in regular practice.

To further investigate the role of choline metabolites in plasma samples as indicators of diet intake, we conducted targeted quantification of choline, betaine, PtdCho, and tHcy, on Day 15 of each arm. Our findings also identified choline and betaine concentrations as the primary markers closely reflecting dietary choline intake. Previous research showed that choline supplementation (930 mg/ day vs 480 mg/day) increases the plasma concentrations of PtdCho in non-pregnant women (47). In our study, plasma PtdCho concentrations did not exhibit a significant association with dietary choline intake and we speculate that 25% AI choline dietary intake for 15 days is not sufficient to reduce the circulating concentration (48–50). Alteration in plasma tHcy concentration had been inversely associated with both choline and betaine dietary intake when folates and vitamin B12 concentrations were low (51), but we did not see this association in our study, where these latter nutrients did not vary between the diet arms.

The use of Fibroscan to assess liver fat content revealed a substantial inter-individual variability, limiting its utility as a stand-alone predictor of choline intake. While a subset of participants exhibited increased liver fat following the 25% AI diet, this response was not consistent with those with the lowest circulating choline concentrations. These observations suggest that factors beyond choline dietary intake, such as dietary factors during the washout period or genetic variability, may influence susceptibility to short term liver fat increase. It is also important to consider that Fibroscan was originally developed for clinical evaluation of liver fibrosis and steatosis and it is optimized for detecting moderate to severe steatosis (52, 53). Moreover, the 15-day duration of low choline intake may not have been sufficient to induce detectable changes by this method. Yet, this merits a study with a more expansive marker evaluation in a more diverse population, to potentially yield more impactful results.

Regression modeling provided additional support for plasma choline and betaine as metabolites that respond to changes in dietary choline intake. In contrast, PtdCho and liver CAP values did not show significant variation. As discussed earlier, this may reflect the relatively short duration for the choline diets or the influence of metabolic compensation mechanisms. Our findings highlight the value of assessing multiple outcomes in parallel to strengthen data interpretation.

The use of a multivariate ROC analysis as a binary classification tool of dietary choline intake showed that choline and betaine together predict accurately dietary choline intake. This approach is cost-effective and accessible after obtaining plasma concentrations of these metabolites and less time consuming compared to traditional tracer studies. Next steps should aim to validate this approach in free-living environments and expand the study to encompass a larger population, including for instance a larger representation of menopausal women, given the well-established influence of hormonal status on dietary choline intake (14, 15).

## CONCLUSION

In summary, our findings demonstrate that plasma choline and betaine are responsive to controlled variations in dietary choline intake, supporting their utility as predictors of dietary intake via ROCs. Importantly, these results underscore the need to expand future research to include genetic variants in choline metabolism and hormonal status as key factors in evaluating individual choline requirements.

## Data Availability

All data produced in the present work are contained in the manuscript

## Conflict of interest

Dr. Steven H. Zeisel has equity in SNP Therapeutics; this paper is not directly related to the company’s product.

Dr. Isis Trujillo-Gonzalez has received research funding from Balchem Corporation, a manufacturer of choline. Dr. Evan M. Paules is a Balchem postdoctoral fellow.

Balchem had no role in the design, analysis or interpretation of the study.

## Data Sharing

Data described in the manuscript will be made available through the University of North Carolina Dataverse repository (URL to be determined).

## Author contributions

ITG: Performed experiments and data analysis, prepared figures and wrote the manuscript. DAH: Conducted data analysis and contributed to manuscript editing. RC and TB: Conducted metabolite measurements. EMP: Performed data analysis and contributed to manuscript editing. EB and WBF: Processed clinical trial samples. AAL and WS: Designed and conducted statistical analysis. MK: Contributed to study design. JS: Coordinated the study and managed participant recruitment. SHZ: Conceived and designed the study, secured funding and contributed to manuscript writing. All authors read and approved the final manuscript.

## Abbreviations

ACho: acetylcholine
AI: adequate intake
ALT: alanine aminotransferase
AST: aspartate aminotransferase
CAP: controlled attenuation parameter
CK: creatine kinase
EREs: estrogen-responsive elements
FDR: false discovery rate
FSH: follicle-stimulating hormone
IER: Isotopic enrichment ratios
MS: mass spectrometry
MTHFR: methylenetetrahydrofolate reductase
PEMT: phosphatidylethanolamine N-methyltransferase
PtdCho: phosphatidylcholine
ROC: receiver operator characteristics
SNPs: single nucleotide polymorphisms
SST: serum-separating tubes
tHcy: total homocysteine

